## Supplementary Appendix for "Factors associated with COVID-19 vaccine uptake in people with kidney disease: an OpenSAFELY cohort study"

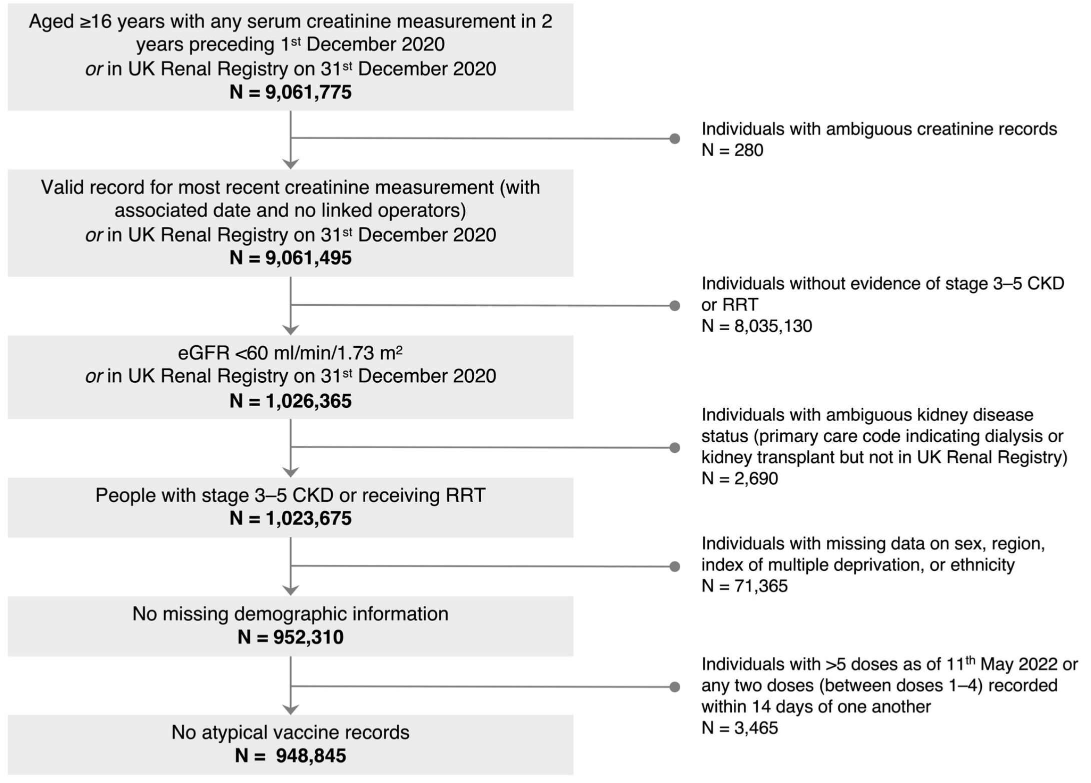


**Supplementary Figure 1. Flow chart of study eligibility.** Frequencies are rounded to the nearest 5. CKD, chronic kidney disease; eGFR, estimated glomerular filtrate rate; RRT, renal replacement therapy.

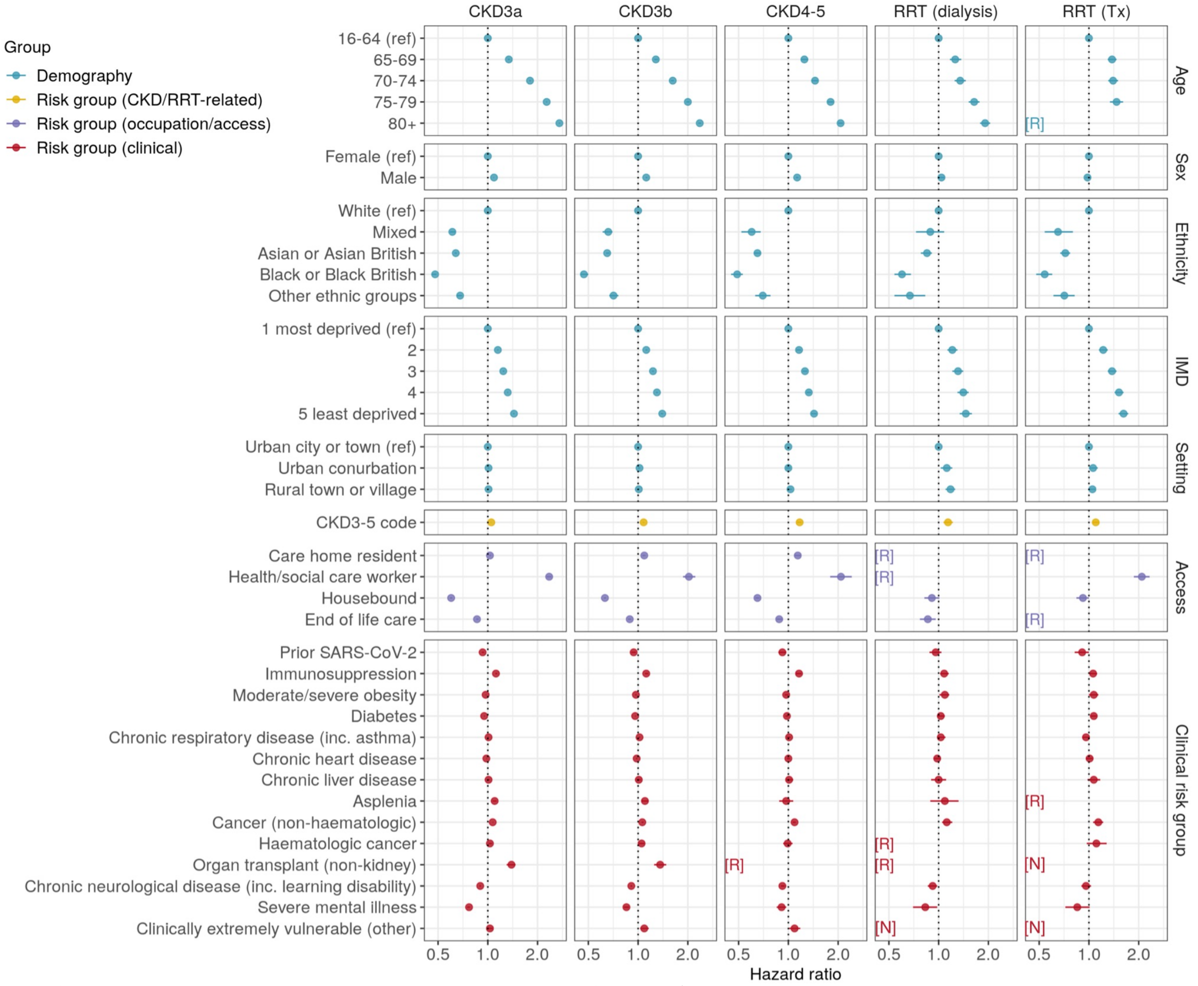


**Supplementary Figure 2. Factors associated with completion of a 4-dose vaccine series in kidney disease subgroups.** Individuals were included in the analysis if they were ≥75 years of age at baseline, care home residents, transplant recipients, or had a history of haematologic malignancy or immunosuppression. Hazard ratios and 95% confidence intervals were derived from partially adjusted models that included age, care home residence, health and social care worker status, housebound status, receipt of end-of-life care, setting (urban/rural), sex, ethnicity, IMD quintile, prior SARS-CoV-2 infection, immunosuppression, and haematologic cancer. Data are redacted [R] for any rows in which there were >0 and ≤10 event or non-event counts after rounding to the nearest 5, with non-event counts calculated among uncensored individuals. CKD, chronic kidney disease; IMD, index of multiple deprivation; [N], covariate absent in all individuals by definition; [R], redacted; RRT, renal replacement therapy.

**Supplementary Table 1. Product profile by dose.**

| Subgroup | Dose 1 | Dose 2 | Dose 3 | Dose 4 |
|---|---|---|---|---|
| **Product** | | | | |
| BNT | 483,280 (53.5%) | 473,820 (53.8%) | 742,985 (92.1%) | 178,065 (47.0%) |
| AZ | 418,795 (46.4%) | 405,975 (46.1%) | 1,910 (0.2%) | 110 (0.0%) |
| MOD | 570 (0.1%) | 905 (0.1%) | 61,515 (7.6%) | 200,445 (52.9%) |
| **2-dose product combination** | | | | |
| BNT-BNT | – | 470,655 (53.4%) | – | – |
| AZ-AZ | – | 402,395 (45.7%) | – | – |
| BNT-AZ | – | 3,580 (0.4%) | – | – |
| AZ-BNT | – | 3,125 (0.4%) | – | – |
| Other | – | 945 (0.1%) | – | – |
| **3-dose product combination** | | | | |
| BNT-BNT-BNT | – | – | 419,020 (52.0%) | – |
| AZ-AZ-BNT | – | – | 320,100 (39.7%) | – |
| AZ-AZ-MOD | – | – | 40,875 (5.1%) | – |
| BNT-BNT-MOD | – | – | 20,015 (2.5%) | – |
| BNT-AZ-BNT | – | – | 2,490 (0.3%) | – |
| AZ-AZ-AZ | – | – | 1,670 (0.2%) | – |
| AZ-BNT-BNT | – | – | 1,185 (0.1%) | – |
| Other | – | – | 1,060 (0.1%) | – |

Frequencies are rounded to the nearest 5. Any dose combinations given to <1000 individuals were pooled as 'Other'. AZ, ChAdOx1-S (AstraZeneca); BNT, BNT162b2 (Pfizer-BioNTech); MOD, mRNA-1273 (Moderna).

**Supplementary Table 2. Cox proportional hazards model and logistic regression outputs for factors associated with completion of a 3-dose vaccination series in people with kidney disease.**

| | Cox models | | | | | Logistic regression | |
|---|---|---|---|---|---|---|---|
| **Variable** | **N (n events)** | **Cov. (%)** | **HR (95% CI), minimally adjusted** | **HR (95% CI), partially adjusted** | **HR (95% CI), fully adjusted** | **N (n events)** | **OR (95% CI), partially adjusted** |
| ***†Age** | | | | | | | |
| 16–64 (ref) | 104,770 (84,815) | 84.1 | 1.00 | 1.00 | 1.00 | 99,200 (83,700) | 1.00 |
| 65–69 | 70,790 (62,075) | 91.6 | 1.32 (1.31–1.33) | 1.29 (1.27–1.30) | 1.31 (1.30–1.33) | 66,485 (61,115) | 1.90 (1.83–1.96) |
| 70–74 | 135,900 (122,380) | 94.5 | 1.75 (1.73–1.77) | 1.67 (1.66–1.69) | 1.71 (1.70–1.73) | 126,665 (120,080) | 2.69 (2.61–2.78) |
| 75–79 | 172,530 (155,085) | 95.5 | 2.18 (2.16–2.20) | 2.09 (2.07–2.11) | 2.15 (2.13–2.17) | 157,535 (150,935) | 3.30 (3.20–3.40) |
| 80+ | 464,860 (381,885) | 95.0 | 2.45 (2.43–2.47) | 2.46 (2.44–2.48) | 2.55 (2.53–2.57) | 371,785 (355,415) | 3.20 (3.12–3.28) |
| **†Sex** | | | | | | | |
| Female (ref) | 530,685 (451,975) | 93.2 | 1.00 | 1.00 | 1.00 | 462,955 (433,405) | 1.00 |
| Male | 418,160 (354,265) | 93.8 | 1.11 (1.11–1.12) | 1.10 (1.09–1.10) | 1.10 (1.10–1.11) | 358,715 (337,840) | 1.18 (1.16–1.20) |
| **†Ethnicity** | | | | | | | |
| White (ref) | 888,875 (764,800) | 94.7 | 1.00 | 1.00 | 1.00 | 768,730 (731,110) | 1.00 |
| Mixed | 5,110 (3,670) | 77.1 | 0.61 (0.59–0.63) | 0.63 (0.61–0.65) | 0.63 (0.61–0.65) | 4,590 (3,560) | 0.30 (0.28–0.32) |
| Asian or Asian British | 32,245 (23,100) | 78.6 | 0.62 (0.61–0.63) | 0.66 (0.65–0.67) | 0.66 (0.65–0.67) | 28,185 (22,340) | 0.33 (0.32–0.35) |
| Black or Black British | 16,345 (10,035) | 65.8 | 0.46 (0.45–0.47) | 0.48 (0.47–0.49) | 0.48 (0.47–0.49) | 14,725 (9,780) | 0.20 (0.20–0.21) |
| Other ethnic groups | 6,270 (4,640) | 81.3 | 0.68 (0.66–0.70) | 0.69 (0.67–0.71) | 0.69 (0.67–0.71) | 5,440 (4,455) | 0.34 (0.31–0.36) |
| **†IMD** | | | | | | | |
| 1 most deprived (ref) | 158,960 (125,290) | 87.6 | 1.00 | 1.00 | 1.00 | 135,750 (119,600) | 1.00 |
| 2 | 180,015 (149,635) | 92.0 | 1.18 (1.17–1.19) | 1.14 (1.14–1.15) | 1.14 (1.13–1.15) | 154,825 (143,160) | 1.44 (1.40–1.48) |
| 3 | 214,280 (184,140) | 94.4 | 1.31 (1.30–1.32) | 1.25 (1.24–1.26) | 1.24 (1.23–1.25) | 185,775 (176,125) | 1.83 (1.78–1.88) |
| 4 | 207,080 (180,565) | 95.5 | 1.40 (1.39–1.41) | 1.32 (1.31–1.33) | 1.31 (1.30–1.32) | 180,325 (172,735) | 2.10 (2.03–2.16) |
| 5 least deprived | 188,510 (166,605) | 96.4 | 1.52 (1.51–1.53) | 1.43 (1.42–1.44) | 1.42 (1.41–1.43) | 164,995 (159,625) | 2.60 (2.52–2.69) |
| **†Setting** | | | | | | | |
| Urban city or town (ref) | 506,660 (432,255) | 93.8 | 1.00 | 1.00 | 1.00 | 439,015 (413,745) | 1 |
| Urban conurbation | 199,215 (160,590) | 89.3 | 0.95 (0.94–0.96) | 1.01 (1.01–1.02) | 1.01 (1.00–1.02) | 170,700 (153,335) | 0.87 (0.85–0.90) |
| Rural town or village | 242,975 (213,390) | 96.0 | 1.08 (1.08–1.09) | 1.01 (1.01–1.02) | 1.01 (1.01–1.02) | 211,955 (204,170) | 1.20 (1.16–1.23) |
| **Kidney disease subgroup** | | | | | | | |
| CKD3a (ref) | 627,265 (547,740) | 93.7 | 1.00 | 1.00 | 1.00 | 563,295 (529,545) | 1.00 |
| CKD3b | 236,205 (194,455) | 93.8 | 0.92 (0.92–0.93) | 0.94 (0.93–0.94) | 0.93 (0.92–0.93) | 193,880 (182,825) | 0.94 (0.92–0.96) |
| CKD4–5 | 63,165 (46,400) | 91.7 | 0.84 (0.83–0.85) | 0.87 (0.87–0.88) | 0.86 (0.85–0.87) | 45,450 (42,055) | 0.84 (0.81–0.88) |
| RRT (dialysis) | 9,440 (6,880) | 86.3 | 1.03 (1.00–1.05) | 1.09 (1.07–1.12) | 1.07 (1.05–1.10) | 7,280 (6,335) | 1.06 (0.99–1.15) |
| RRT (transplant) | 12,770 (10,765) | 88.7 | 1.54 (1.51–1.57) | 1.45 (1.42–1.49) | 1.44 (1.41–1.47) | 11,765 (10,490) | 1.09 (1.01–1.17) |
| **Primary care coding of kidney disease** | | | | | | | |
| CKD diagnostic code | 563,335 (478,860) | 94.1 | 1.03 (1.02–1.03) | 1.04 (1.03–1.04) | 1.06 (1.06–1.07) | 482,525 (456,065) | 1.23 (1.21–1.25) |
| **Risk group (occupation/access)** | | | | | | | |
| *†Care home resident | 43,385 (27,150) | 94.5 | 1.01 (1.00–1.02) | 1.06 (1.05–1.07) | 1.11 (1.10–1.12) | 23,070 (21,910) | 1.02 (0.96–1.09) |
| *†Health/social care worker | 5,175 (4,770) | 93.2 | 2.15 (2.09–2.21) | 2.27 (2.21–2.34) | 2.25 (2.19–2.32) | 5,060 (4,730) | 2.66 (2.36–2.99) |
| †Housebound | 53,520 (37,175) | 92.4 | 0.60 (0.59–0.60) | 0.61 (0.61–0.62) | 0.63 (0.62–0.63) | 35,160 (32,795) | 0.79 (0.76–0.83) |

| | | | | | | | |
|---|---|---|---|---|---|---|---|
| †End of life care | 39,430 (23,495) | 92.6 | 0.82 (0.81–0.83) | 0.87 (0.85–0.88) | 0.87 (0.86–0.88) | 21,080 (19,740) | 0.86 (0.81–0.91) |
| **Risk group (clinical)** | | | | | | | |
| †Prior SARS-CoV-2 | 25,490 (17,785) | 90.7 | 0.89 (0.88–0.91) | 0.94 (0.92–0.95) | 0.94 (0.93–0.96) | 17,785 (16,230) | 0.96 (0.91–1.02) |
| †Immunosuppression | 80,025 (67,140) | 93.8 | 1.17 (1.16–1.18) | 1.19 (1.18–1.20) | 1.11 (1.10–1.12) | 67,605 (63,680) | 1.39 (1.34–1.45) |
| Moderate/severe obesity | 108,950 (94,075) | 92.5 | 0.93 (0.92–0.94) | 0.97 (0.96–0.97) | 0.98 (0.97–0.99) | 98,525 (91,440) | 1.11 (1.08–1.14) |
| Diabetes | 269,895 (222,880) | 92.6 | 0.91 (0.90–0.91) | 0.95 (0.95–0.96) | 0.96 (0.96–0.97) | 227,180 (211,585) | 1.06 (1.04–1.08) |
| CRD (inc. asthma) | 210,095 (174,610) | 93.5 | 0.98 (0.98–0.99) | 1.01 (1.00–1.01) | 1.01 (1.01–1.02) | 176,195 (165,520) | 1.05 (1.03–1.08) |
| Chronic heart disease | 425,575 (352,775) | 94.4 | 0.98 (0.98–0.99) | 0.98 (0.97–0.98) | 0.99 (0.98–0.99) | 349,835 (331,995) | 1.06 (1.04–1.09) |
| Chronic liver disease | 31,285 (26,060) | 91.9 | 1.00 (0.99–1.01) | 1.01 (0.99–1.02) | 1.01 (1.00–1.02) | 27,045 (24,990) | 1.01 (0.96–1.06) |
| Asplenia | 8,405 (7,180) | 94.3 | 1.10 (1.07–1.12) | 1.08 (1.06–1.11) | 1.08 (1.06–1.11) | 7,270 (6,875) | 1.19 (1.07–1.32) |
| Cancer (non-haematologic) | 164,110 (136,440) | 95.4 | 1.09 (1.08–1.10) | 1.07 (1.06–1.08) | 1.07 (1.06–1.08) | 134,180 (128,730) | 1.30 (1.27–1.34) |
| †Haematologic cancer | 19,990 (16,325) | 95.4 | 1.16 (1.14–1.18) | 0.99 (0.97–1.00) | 1.04 (1.03–1.06) | 15,800 (15,175) | 1.07 (0.98–1.18) |
| Organ transplant (non-kidney) | 2,170 (1,865) | 93.0 | 1.46 (1.39–1.52) | 1.31 (1.26–1.38) | 1.39 (1.33–1.46) | 1,910 (1,795) | 1.41 (1.16–1.72) |
| CND (inc. learning disability) | 158,200 (124,130) | 93.4 | 0.89 (0.88–0.89) | 0.90 (0.90–0.91) | 0.91 (0.90–0.91) | 121,505 (114,275) | 0.90 (0.87–0.92) |
| Severe mental illness | 16,270 (12,665) | 87.2 | 0.76 (0.75–0.77) | 0.79 (0.78–0.81) | 0.80 (0.78–0.81) | 13,735 (12,055) | 0.67 (0.63–0.70) |
| CEV (other) | 5,750 (4,815) | 91.6 | 1.01 (0.98–1.03) | 1.05 (1.02–1.08) | 1.03 (1.00–1.06) | 4,965 (4,580) | 0.93 (0.83–1.04) |

Cumulative coverage was determined based on Kaplan-Meier estimates, censoring on death, deregistration, or 11$^{th}$ May 2022. Logistic regression models included 821,670 individuals who were uncensored (registered and alive) as of the analysis cut-off (11$^{th}$ May 2022). * variable included in all minimally adjusted models; † variable included in all partially adjusted models. CI, confidence interval; CKD, chronic kidney disease; HR, hazard ratio; IMD, index of multiple deprivation; OR, odds ratio; RRT, renal replacement therapy.

**Supplementary Table 3. Cox proportional hazards models for factors associated with completion of a 3-dose vaccination series in kidney disease subgroups.**

| | CKD3a | | | CKD3b | | | CKD4–5 | | | RRT (dialysis) | | | RRT (transplant) | | |
|---|---|---|---|---|---|---|---|---|---|---|---|---|---|---|---|
| Variable | N (n events) | Cov. (%) | HR (95% CI) | N (n events) | Cov. (%) | HR (95% CI) | N (n events) | Cov. (%) | HR (95% CI) | N (n events) | Cov. (%) | HR (95% CI) | N (n events) | Cov. (%) | HR (95% CI) |
| ***†Age** | | | | | | | | | | | | | | | |
| 16–64 (ref) | 71,970 (59,135) | 84.5 | 1.00 | 13,245 (10,360) | 82.5 | 1.00 | 5,870 (4,385) | 80.7 | 1.00 | 4,390 (3,165) | 79.9 | 1.00 | 9,295 (7,770) | 86.7 | 1.00 |
| 65–69 | 54,435 (48,425) | 92.0 | 1.34 (1.32–1.36) | 10,815 (9,205) | 90.5 | 1.28 (1.25–1.32) | 3,020 (2,355) | 88.1 | 1.25 (1.19–1.32) | 1,095 (830) | 88.0 | 1.26 (1.17–1.37) | 1,425 (1,260) | 94.3 | 1.38 (1.30–1.47) |
| 70–74 | 102,975 (94,125) | 94.8 | 1.80 (1.78–1.82) | 24,730 (21,670) | 93.8 | 1.62 (1.58–1.65) | 5,865 (4,700) | 91.2 | 1.45 (1.39–1.51) | 1,145 (855) | 90.3 | 1.35 (1.25–1.46) | 1,180 (1,035) | 94.2 | 1.40 (1.31–1.50) |
| 75–79 | 124,030 (113,350) | 95.7 | 2.27 (2.25–2.29) | 38,245 (33,655) | 95.0 | 2.00 (1.96–2.05) | 8,410 (6,645) | 93.5 | 1.80 (1.73–1.87) | 1,220 (920) | 94.5 | 1.64 (1.52–1.77) | 620 (510) | 94.0 | 1.47 (1.34–1.61) |
| 80+ | 273,855 (232,705) | 95.3 | 2.71 (2.69–2.74) | 149,170 (119,565) | 94.8 | 2.36 (2.31–2.41) | 40,000 (28,315) | 93.5 | 2.07 (2.00–2.14) | 1,590 (1,110) | 93.1 | 1.91 (1.78–2.05) | [R] | [R] | [R] |
| **†Sex** | | | | | | | | | | | | | | | |
| Female (ref) | 355,460 (310,735) | 93.4 | 1.00 | 134,120 (110,490) | 93.4 | 1.00 | 32,700 (24,050) | 91.3 | 1.00 | 3,480 (2,530) | 84.7 | 1.00 | 4,925 (4,175) | 88.5 | 1.00 |
| Male | 271,805 (237,005) | 94.0 | 1.09 (1.08–1.09) | 102,085 (83,965) | 94.2 | 1.12 (1.11–1.14) | 30,465 (22,355) | 92.1 | 1.13 (1.11–1.15) | 5,960 (4,355) | 87.0 | 1.04 (0.99–1.09) | 7,845 (6,590) | 88.6 | 0.98 (0.95–1.02) |
| **†Ethnicity** | | | | | | | | | | | | | | | |
| White (ref) | 590,315 (521,735) | 94.9 | 1.00 | 222,900 (185,385) | 94.9 | 1.00 | 57,975 (43,250) | 93.5 | 1.00 | 7,295 (5,415) | 88.9 | 1.00 | 10,390 (9,015) | 91.2 | 1.00 |
| Mixed | 3,415 (2,470) | 76.1 | 0.61 (0.59–0.64) | 1,065 (770) | 79.8 | 0.66 (0.61–0.70) | 350 (220) | 73.6 | 0.60 (0.52–0.68) | 140 (105) | 76.9 | 0.89 (0.73–1.08) | 140 (100) | 71.4 | 0.65 (0.54–0.80) |
| Asian or Asian British | 18,720 (13,805) | 79.0 | 0.64 (0.63–0.65) | 7,595 (5,310) | 78.3 | 0.65 (0.64–0.67) | 3,120 (1,915) | 74.7 | 0.65 (0.62–0.68) | 1,295 (920) | 80.6 | 0.85 (0.78–0.91) | 1,510 (1,145) | 79.4 | 0.72 (0.67–0.77) |
| Black or Black British | 10,840 (6,710) | 65.1 | 0.48 (0.46–0.49) | 3,240 (1,965) | 66.8 | 0.47 (0.45–0.50) | 1,225 (695) | 65.5 | 0.49 (0.45–0.53) | 570 (355) | 67.5 | 0.60 (0.54–0.68) | 470 (310) | 68.9 | 0.54 (0.48–0.60) |
| Other ethnic groups | 3,975 (3,015) | 81.6 | 0.68 (0.65–0.70) | 1,405 (1,025) | 82.0 | 0.71 (0.67–0.76) | 495 (320) | 76.1 | 0.70 (0.63–0.78) | 135 (85) | 66.2 | 0.67 (0.54–0.83) | 260 (190) | 76.7 | 0.71 (0.61–0.82) |
| **†IMD** | | | | | | | | | | | | | | | |
| 1 most deprived (ref) | 100,965 (82,025) | 87.9 | 1.00 | 40,770 (31,390) | 88.5 | 1.00 | 12,015 (8,120) | 84.9 | 1.00 | 2,610 (1,770) | 78.8 | 1.00 | 2,600 (1,990) | 80.2 | 1.00 |
| 2 | 116,850 (99,945) | 92.2 | 1.15 (1.14–1.16) | 45,630 (36,860) | 92.5 | 1.12 (1.11–1.14) | 12,685 (9,090) | 90.2 | 1.16 (1.12–1.19) | 2,185 (1,555) | 84.8 | 1.21 (1.13–1.30) | 2,665 (2,185) | 86.0 | 1.22 (1.15–1.30) |
| 3 | 141,655 (124,995) | 94.6 | 1.24 (1.23–1.26) | 53,770 (44,755) | 94.8 | 1.23 (1.21–1.25) | 14,205 (10,595) | 92.9 | 1.26 (1.22–1.30) | 1,890 (1,425) | 88.2 | 1.31 (1.21–1.41) | 2,760 (2,370) | 90.2 | 1.38 (1.30–1.47) |
| 4 | 139,220 (124,375) | 95.6 | 1.32 (1.31–1.33) | 50,790 (42,810) | 95.6 | 1.30 (1.28–1.32) | 13,015 (9,975) | 94.5 | 1.33 (1.29–1.37) | 1,530 (1,195) | 91.2 | 1.41 (1.30–1.52) | 2,520 (2,210) | 92.2 | 1.52 (1.43–1.62) |
| 5 least deprived | 128,575 (116,400) | 96.6 | 1.44 (1.42–1.45) | 45,240 (38,635) | 96.4 | 1.40 (1.37–1.42) | 11,245 (8,625) | 95.4 | 1.43 (1.38–1.48) | 1,230 (935) | 91.6 | 1.46 (1.34–1.59) | 2,225 (2,010) | 93.8 | 1.62 (1.51–1.73) |
| **†Setting** | | | | | | | | | | | | | | | |
| Urban city or town (ref) | 334,925 (293,565) | 94.0 | 1.00 | 126,760 (104,820) | 94.2 | 1.00 | 33,625 (24,845) | 92.4 | 1.00 | 4,735 (3,400) | 86.0 | 1.00 | 6,615 (5,630) | 89.3 | 1.00 |
| Urban conurbation | 128,630 (106,790) | 89.7 | 1.01 (1.00–1.02) | 49,545 (38,800) | 89.8 | 1.02 (1.00–1.03) | 14,540 (10,095) | 86.3 | 1.00 (0.97–1.03) | 3,000 (2,135) | 82.4 | 1.12 (1.03–1.21) | 3,495 (2,770) | 83.1 | 1.06 (1.00–1.13) |
| Rural town or village | 163,705 (147,385) | 96.1 | 1.01 (1.00–1.02) | 59,900 (50,830) | 96.1 | 1.01 (1.00–1.02) | 15,000 (11,460) | 95.2 | 1.03 (1.01–1.05) | 1,705 (1,345) | 93.2 | 1.18 (1.10–1.26) | 2,660 (2,370) | 93.5 | 1.05 (1.00–1.10) |
| **Primary care coding of kidney disease** | | | | | | | | | | | | | | | |
| CKD diagnostic code | 306,820 (270,530) | 94.5 | 1.05 (1.04–1.05) | 183,225 (152,815) | 94.3 | 1.08 (1.07–1.09) | 55,645 (41,425) | 92.2 | 1.17 (1.13–1.20) | 7,990 (5,895) | 87.6 | 1.14 (1.07–1.22) | 9,655 (8,200) | 89.6 | 1.10 (1.05–1.15) |
| **Risk group (occupation/access)** | | | | | | | | | | | | | | | |
| *†Care home resident | 23,700 (15,635) | 94.5 | 1.03 (1.02–1.05) | 14,845 (8,955) | 94.6 | 1.09 (1.06–1.11) | 4,605 (2,425) | 93.2 | 1.14 (1.09–1.19) | [R] | [R] | [R] | [R] | [R] | [R] |
| *†Health/social care worker | 3,975 (3,680) | 93.5 | 2.35 (2.27–2.43) | 585 (530) | 89.7 | 2.03 (1.87–2.22) | 195 (170) | 84.2 | 2.08 (1.79–2.42) | [R] | [R] | [R] | 355 (335) | 91.7 | 2.09 (1.87–2.33) |
| †Housebound | 26,735 (19,485) | 92.7 | 0.60 (0.59–0.61) | 18,770 (12,815) | 92.4 | 0.63 (0.62–0.64) | 6,800 (3,985) | 90.4 | 0.65 (0.63–0.67) | 600 (370) | 88.4 | 0.91 (0.82–1.01) | 615 (520) | 89.2 | 0.92 (0.84–1.00) |
| †End of life care | 20,070 (12,665) | 92.9 | 0.86 (0.85–0.88) | 13,185 (7,700) | 92.9 | 0.89 (0.87–0.91) | 5,385 (2,640) | 90.7 | 0.88 (0.85–0.92) | 600 (355) | 84.7 | 0.86 (0.77–0.96) | [R] | [R] | [R] |
| **Risk group (clinical)** | | | | | | | | | | | | | | | |
| †Prior SARS-CoV-2 | 14,290 (10,625) | 91.4 | 0.93 (0.92–0.95) | 7,135 (4,690) | 91.5 | 0.94 (0.91–0.97) | 2,545 (1,400) | 88.1 | 0.92 (0.87–0.97) | 1,000 (670) | 82.0 | 0.96 (0.88–1.04) | 515 (400) | 81.5 | 0.91 (0.82–1.00) |
| Immunosuppression | 43,575 (37,735) | 94.9 | 1.12 (1.11–1.14) | 18,245 (14,895) | 94.8 | 1.12 (1.10–1.15) | 5,525 (3,980) | 92.7 | 1.16 (1.11–1.20) | 2,065 (1,540) | 85.6 | 1.08 (1.02–1.15) | 10,615 (8,995) | 89.1 | 1.06 (1.01–1.12) |
| Moderate/severe obesity | 71,070 (62,710) | 92.6 | 0.97 (0.97–0.98) | 27,390 (23,215) | 92.9 | 0.97 (0.96–0.98) | 8,000 (6,145) | 90.7 | 0.97 (0.95–1.00) | 1,325 (1,005) | 87.5 | 1.09 (1.01–1.16) | 1,160 (995) | 89.0 | 1.07 (1.00–1.14) |
| Diabetes | 152,745 (130,485) | 92.8 | 0.95 (0.95–0.96) | 81,390 (66,145) | 92.9 | 0.96 (0.96–0.97) | 27,680 (20,000) | 91.0 | 0.98 (0.96–1.00) | 4,040 (2,860) | 87.5 | 1.03 (0.98–1.09) | 4,040 (3,390) | 89.9 | 1.07 (1.02–1.11) |
| CRD (inc. asthma) | 136,395 (116,745) | 93.6 | 1.01 (1.00–1.01) | 54,735 (44,070) | 93.9 | 1.02 (1.01–1.03) | 14,675 (10,480) | 91.7 | 1.01 (0.98–1.03) | 2,035 (1,440) | 86.9 | 1.03 (0.98–1.10) | 2,255 (1,880) | 88.0 | 0.96 (0.91–1.01) |
| Chronic heart disease | 255,835 (219,880) | 94.7 | 0.98 (0.98–0.99) | 125,250 (100,865) | 94.4 | 0.98 (0.97–0.99) | 35,895 (25,480) | 92.8 | 1.00 (0.98–1.02) | 4,740 (3,355) | 87.7 | 0.98 (0.94–1.03) | 3,860 (3,195) | 90.1 | 1.01 (0.96–1.05) |
| Chronic liver disease | 20,250 (17,365) | 92.1 | 1.01 (1.00–1.03) | 7,650 (6,190) | 92.3 | 1.01 (0.98–1.04) | 2,245 (1,630) | 90.3 | 1.01 (0.97–1.07) | 550 (380) | 80.5 | 1.00 (0.90–1.11) | 585 (490) | 88.8 | 1.07 (0.98–1.17) |
| Asplenia | 5,485 (4,835) | 94.7 | 1.10 (1.07–1.13) | 2,035 (1,695) | 93.5 | 1.10 (1.05–1.15) | 575 (400) | 87.4 | 0.97 (0.88–1.07) | 140 (100) | 83.3 | 1.09 (0.89–1.32) | [R] | [R] | [R] |
| Cancer (non-haematologic) | 103,955 (89,290) | 95.6 | 1.07 (1.07–1.08) | 45,090 (36,250) | 95.5 | 1.06 (1.05–1.08) | 12,800 (9,115) | 94.1 | 1.09 (1.06–1.11) | 1,305 (965) | 91.4 | 1.12 (1.05–1.21) | 960 (820) | 92.0 | 1.14 (1.06–1.22) |
| Haematologic cancer | 11,885 (10,075) | 95.6 | 1.03 (1.01–1.06) | 5,705 (4,560) | 95.5 | 1.05 (1.01–1.09) | 1,845 (1,265) | 92.7 | 0.99 (0.93–1.06) | [R] | [R] | [R] | 240 (205) | 89.6 | 1.11 (0.97–1.28) |
| Organ transplant (non-kidney) | 1,195 (1,055) | 92.0 | 1.39 (1.30–1.47) | 685 (575) | 93.0 | 1.36 (1.25–1.48) | [R] | [R] | [R] | [R] | [R] | [R] | [N] | [N] | [N] |
| CND (inc. learning disability) | 96,415 (78,640) | 93.7 | 0.90 (0.89–0.91) | 46,255 (34,965) | 93.4 | 0.91 (0.90–0.92) | 12,905 (8,540) | 91.6 | 0.92 (0.90–0.95) | 1,440 (995) | 86.0 | 0.92 (0.86–0.98) | 1,185 (985) | 89.2 | 0.96 (0.90–1.03) |
| Severe mental illness | 10,465 (8,255) | 86.4 | 0.77 (0.75–0.78) | 4,105 (3,205) | 89.6 | 0.85 (0.82–0.88) | 1,310 (925) | 87.1 | 0.91 (0.85–0.97) | 220 (145) | 72.2 | 0.83 (0.70–0.98) | 170 (135) | 81.2 | 0.85 (0.72–1.01) |
| CEV (other) | 3,480 (2,990) | 92.1 | 1.03 (0.99–1.07) | 1,465 (1,195) | 92.1 | 1.09 (1.03–1.16) | 810 (630) | 87.3 | 1.09 (1.01–1.18) | [N] | [N] | [N] | [N] | [N] | [N] |

Counts are rounded to the nearest 5. Cumulative coverage was determined based on Kaplan-Meier estimates, censoring on death, deregistration, or 11th May 2022. Hazard ratios and 95% confidence intervals were derived from partially adjusted models that included age, care home residence, health and social care worker status, housebound status, receipt of end-of-life care, setting (urban/rural), sex, ethnicity, IMD quintile, prior SARS-CoV-2 infection, immunosuppression, and haematologic cancer. Data are redacted [R] for any rows in which there were >0 and ≤10 event or non-event counts after rounding to the nearest 5, with non-event counts calculated among uncensored individuals. CEV, clinically extremely vulnerable; CKD, chronic kidney disease; CND, chronic neurological disease; IMD, index of multiple deprivation; [N], covariate absent in all individuals by definition; [R] redacted; RRT, renal replacement therapy.

**Supplementary Table 4. Baseline characteristics of secondary outcome analysis population (dose 4 uptake).**

| Characteristic | All<br>N = 679,405 | CKD3a<br>N = 418,095 | CKD3b<br>N = 193,475 | CKD4–5<br>N = 50,465 | RRT (dialysis)<br>N = 4,595 | RRT (transplant)<br>N = 12,770 |
|---|---|---|---|---|---|---|
| **Age** | | | | | | |
| 16–64 | 20,015 (2.9%) | 6,775 (1.6%) | 1,845 (1.0%) | 800 (1.6%) | 1,300 (28.3%) | 9,295 (72.8%) |
| 65–69 | 8,210 (1.2%) | 4,705 (1.1%) | 1,395 (0.7%) | 435 (0.9%) | 245 (5.3%) | 1,425 (11.2%) |
| 70–74 | 13,795 (2.0%) | 8,730 (2.1%) | 2,815 (1.5%) | 825 (1.6%) | 245 (5.3%) | 1,180 (9.2%) |
| 75–79 | 172,530 (25.4%) | 124,030 (29.7%) | 38,245 (19.8%) | 8,410 (16.7%) | 1,220 (26.6%) | 620 (4.9%) |
| 80+ | 464,860 (68.4%) | 273,855 (65.5%) | 149,170 (77.1%) | 40,000 (79.3%) | 1,590 (34.6%) | 250 (2.0%) |
| **Sex** | | | | | | |
| Female | 387,500 (57.0%) | 241,015 (57.6%) | 112,810 (58.3%) | 27,035 (53.6%) | 1,710 (37.2%) | 4,925 (38.6%) |
| Male | 291,905 (43.0%) | 177,080 (42.4%) | 80,665 (41.7%) | 23,430 (46.4%) | 2,885 (62.8%) | 7,845 (61.4%) |
| **Ethnicity** | | | | | | |
| White | 645,830 (95.1%) | 399,635 (95.6%) | 184,715 (95.5%) | 47,335 (93.8%) | 3,755 (81.7%) | 10,390 (81.4%) |
| Mixed | 2,740 (0.4%) | 1,620 (0.4%) | 710 (0.4%) | 215 (0.4%) | 60 (1.3%) | 140 (1.1%) |
| Asian or Asian British | 18,695 (2.8%) | 9,955 (2.4%) | 4,920 (2.5%) | 1,820 (3.6%) | 490 (10.7%) | 1,510 (11.8%) |
| Black or Black British | 8,275 (1.2%) | 4,630 (1.1%) | 2,155 (1.1%) | 780 (1.5%) | 235 (5.1%) | 470 (3.7%) |
| Other ethnic groups | 3,865 (0.6%) | 2,255 (0.5%) | 980 (0.5%) | 315 (0.6%) | 55 (1.2%) | 260 (2.0%) |
| **Index of multiple deprivation quintile** | | | | | | |
| 1 most deprived | 103,695 (15.3%) | 60,420 (14.5%) | 30,950 (16.0%) | 8,705 (17.2%) | 1,015 (22.1%) | 2,600 (20.4%) |
| 2 | 124,970 (18.4%) | 74,870 (17.9%) | 36,495 (18.9%) | 9,935 (19.7%) | 1,005 (21.9%) | 2,665 (20.9%) |
| 3 | 155,525 (22.9%) | 95,730 (22.9%) | 44,525 (23.0%) | 11,530 (22.8%) | 980 (21.3%) | 2,760 (21.6%) |
| 4 | 152,900 (22.5%) | 95,975 (23.0%) | 42,745 (22.1%) | 10,810 (21.4%) | 845 (18.4%) | 2,520 (19.7%) |
| 5 least deprived | 142,315 (20.9%) | 91,100 (21.8%) | 38,755 (20.0%) | 9,485 (18.8%) | 745 (16.2%) | 2,225 (17.4%) |
| **Setting** | | | | | | |
| Urban city or town | 362,390 (53.3%) | 222,365 (53.2%) | 103,970 (53.7%) | 27,115 (53.7%) | 2,320 (50.5%) | 6,615 (51.8%) |
| Urban conurbation | 138,875 (20.4%) | 83,815 (20.0%) | 39,355 (20.3%) | 10,895 (21.6%) | 1,320 (28.7%) | 3,495 (27.4%) |
| Rural | 178,145 (26.2%) | 111,915 (26.8%) | 50,150 (25.9%) | 12,460 (24.7%) | 960 (20.9%) | 2,660 (20.8%) |
| **Primary care coding of kidney disease** | | | | | | |
| CKD3–5 diagnostic code | 424,480 (62.5%) | 214,160 (51.2%) | 151,790 (78.5%) | 44,890 (89.0%) | 3,985 (86.7%) | 9,655 (75.6%) |
| Dialysis code | 12,025 (1.8%) | 0 (0.0%) | 0 (0.0%) | 0 (0.0%) | 3,720 (81.0%) | 8,305 (65.0%) |
| Kidney transplant code | 13,435 (2.0%) | 0 (0.0%) | 0 (0.0%) | 0 (0.0%) | 1,070 (23.3%) | 12,360 (96.8%) |
| **Risk group (occupation/access)** | | | | | | |
| Care home resident | 43,385 (6.4%) | 23,700 (5.7%) | 14,845 (7.7%) | 4,605 (9.1%) | 165 (3.6%) | 65 (0.5%) |
| Health/social care worker | 1,095 (0.2%) | 565 (0.1%) | 120 (0.1%) | 35 (0.1%) | 20 (0.4%) | 355 (2.8%) |
| Housebound | 49,420 (7.3%) | 24,345 (5.8%) | 17,740 (9.2%) | 6,345 (12.6%) | 375 (8.2%) | 615 (4.8%) |
| End of life care | 35,765 (5.3%) | 17,935 (4.3%) | 12,290 (6.4%) | 4,995 (9.9%) | 365 (7.9%) | 180 (1.4%) |
| **Risk group (clinical)** | | | | | | |
| Prior SARS-CoV-2 * | 19,170 (2.8%) | 10,080 (2.4%) | 6,040 (3.1%) | 2,065 (4.1%) | 475 (10.3%) | 515 (4.0%) |
| Immunosuppression | 80,025 (11.8%) | 43,575 (10.4%) | 18,245 (9.4%) | 5,525 (10.9%) | 2,065 (44.9%) | 10,615 (83.1%) |
| Moderate/severe obesity | 55,040 (8.1%) | 31,670 (7.6%) | 16,970 (8.8%) | 4,880 (9.7%) | 360 (7.8%) | 1,160 (9.1%) |
| Diabetes | 190,140 (28.0%) | 100,110 (23.9%) | 62,990 (32.6%) | 21,315 (42.2%) | 1,685 (36.7%) | 4,040 (31.6%) |
| Chronic respiratory disease (inc. asthma) | 151,400 (22.3%) | 91,725 (21.9%) | 44,605 (23.1%) | 11,810 (23.4%) | 1,005 (21.9%) | 2,255 (17.7%) |
| Chronic heart disease | 340,605 (50.1%) | 195,385 (46.7%) | 107,965 (55.8%) | 30,860 (61.2%) | 2,535 (55.2%) | 3,860 (30.2%) |

| | | | | | | |
|---|---|---|---|---|---|---|
| Chronic liver disease | 18,715 (2.8%) | 11,090 (2.7%) | 5,290 (2.7%) | 1,505 (3.0%) | 240 (5.2%) | 585 (4.6%) |
| Asplenia | 5,930 (0.9%) | 3,650 (0.9%) | 1,575 (0.8%) | 455 (0.9%) | 80 (1.7%) | 170 (1.3%) |
| Cancer (non-haematologic) | 128,475 (18.9%) | 77,375 (18.5%) | 38,315 (19.8%) | 11,020 (21.8%) | 805 (17.5%) | 960 (7.5%) |
| Haematologic cancer | 19,990 (2.9%) | 11,885 (2.8%) | 5,705 (2.9%) | 1,845 (3.7%) | 310 (6.7%) | 240 (1.9%) |
| Organ transplant (non-kidney) † | 2,170 (0.3%) | 1,195 (0.3%) | 685 (0.4%) | 215 (0.4%) | 75 (1.6%) | [N] |
| CND (inc. learning disability) | 131,900 (19.4%) | 77,535 (18.5%) | 41,125 (21.3%) | 11,295 (22.4%) | 765 (16.6%) | 1,185 (9.3%) |
| Severe mental illness | 9,190 (1.4%) | 5,405 (1.3%) | 2,700 (1.4%) | 835 (1.7%) | 80 (1.7%) | 170 (1.3%) |
| Clinically extremely vulnerable (other) § | 3,990 (0.6%) | 2,370 (0.6%) | 1,170 (0.6%) | 450 (0.9%) | [N] | [N] |
| **Region** | | | | | | |
| London | 20,255 (3.0%) | 11,740 (2.8%) | 5,490 (2.8%) | 1,800 (3.6%) | 315 (6.9%) | 910 (7.1%) |
| East of England | 152,635 (22.5%) | 94,895 (22.7%) | 42,880 (22.2%) | 10,840 (21.5%) | 1,055 (23.0%) | 2,970 (23.3%) |
| East Midlands | 125,850 (18.5%) | 77,615 (18.6%) | 35,975 (18.6%) | 9,215 (18.3%) | 790 (17.2%) | 2,250 (17.6%) |
| North East | 31,955 (4.7%) | 19,750 (4.7%) | 9,150 (4.7%) | 2,215 (4.4%) | 240 (5.2%) | 595 (4.7%) |
| North West | 62,430 (9.2%) | 38,315 (9.2%) | 17,940 (9.3%) | 4,700 (9.3%) | 305 (6.6%) | 1,165 (9.1%) |
| South East | 42,580 (6.3%) | 26,645 (6.4%) | 11,810 (6.1%) | 3,100 (6.1%) | 290 (6.3%) | 735 (5.8%) |
| South West | 122,945 (18.1%) | 76,210 (18.2%) | 35,105 (18.1%) | 9,145 (18.1%) | 695 (15.1%) | 1,790 (14.0%) |
| West Midlands | 24,805 (3.7%) | 14,830 (3.5%) | 7,255 (3.7%) | 1,975 (3.9%) | 275 (6.0%) | 465 (3.6%) |
| Yorkshire and the Humber | 95,950 (14.1%) | 58,095 (13.9%) | 27,870 (14.4%) | 7,480 (14.8%) | 620 (13.5%) | 1,885 (14.8%) |

Individuals were included in the analysis if they were ≥75 years of age at baseline, care home residents, transplant recipients, or had a history of haematologic malignancy or immunosuppression. Data are n (%) after rounding to the nearest 5. Primary care codes and risk groups are coded by separate binary variables; percentages under these table subheadings therefore do not sum to 100. Among people with CKD4–5, 46,830 had stage 4 CKD (eGFR 15–29 ml/min/1.73 $m^2$) and 3,635 had stage 5 CKD (eGFR <15ml/min/1.73 $m^2$). CKD, chronic kidney disease; CND, chronic neurological disease; [N], covariate absent in all individuals by definition; RRT, renal replacement therapy. * Based on prior evidence of a positive SARS-CoV-2 test, COVID-19-related primary care code, or COVID-19-related hospitalisation as of 1st December 2020; † Excludes individuals with kidney transplants based on primary care coding or UK Renal Registry status; § Classified as clinically extremely vulnerable in the absence of any of the comorbidities listed above (including RRT).

**Supplementary Table 5. Cox proportional hazards models for factors associated with completion of a 4-dose vaccination series in people with kidney disease.**

| Variable | N (n events) | Cov. (%) | HR (95% CI), minimally adjusted | HR (95% CI), partially adjusted | HR (95% CI), fully adjusted |
|---|---|---|---|---|---|
| ***†Age** | | | | | |
| 16–64 (ref) | 20,015 (8,005) | 42.5 | 1.00 | 1.00 | 1.00 |
| 65–69 | 8,210 (3,340) | 44.9 | 1.08 (1.04-1.12) | 0.99 (0.95-1.03) | 1.52 (1.45-1.58) |
| 70–74 | 13,795 (6,090) | 50.0 | 1.22 (1.18-1.26) | 1.06 (1.03-1.10) | 1.80 (1.73-1.87) |
| 75–79 | 172,530 (99,315) | 62.8 | 1.38 (1.35-1.41) | 1.51 (1.47-1.54) | 2.76 (2.67-2.85) |
| 80+ | 464,860 (232,695) | 61.9 | 1.41 (1.38-1.45) | 1.59 (1.55-1.64) | 2.95 (2.86-3.05) |
| **†Sex** | | | | | |
| Female (ref) | 387,500 (192,935) | 58.6 | 1.00 | 1.00 | 1.00 |
| Male | 291,905 (156,510) | 64.4 | 1.19 (1.18-1.19) | 1.17 (1.16-1.17) | 1.16 (1.16-1.17) |
| **†Ethnicity** | | | | | |
| White (ref) | 645,830 (340,050) | 62.5 | 1.00 | 1.00 | 1.00 |
| Mixed | 2,740 (875) | 36.9 | 0.51 (0.47-0.54) | 0.53 (0.50-0.57) | 0.54 (0.50-0.57) |
| Asian or Asian British | 18,695 (5,205) | 32.8 | 0.44 (0.43-0.45) | 0.48 (0.46-0.49) | 0.47 (0.45-0.48) |
| Black or Black British | 8,275 (1,805) | 25.1 | 0.32 (0.31-0.34) | 0.35 (0.33-0.37) | 0.36 (0.34-0.37) |
| Other ethnic groups | 3,865 (1,510) | 46.5 | 0.69 (0.66-0.73) | 0.71 (0.68-0.75) | 0.69 (0.66-0.73) |
| **†IMD** | | | | | |
| 1 most deprived (ref) | 103,695 (41,680) | 48.9 | 1.00 | 1.00 | 1.00 |
| 2 | 124,970 (58,805) | 56.5 | 1.23 (1.22-1.25) | 1.19 (1.18-1.21) | 1.19 (1.17-1.20) |
| 3 | 155,525 (81,110) | 61.8 | 1.40 (1.38-1.41) | 1.32 (1.30-1.34) | 1.31 (1.29-1.33) |
| 4 | 152,900 (84,305) | 64.8 | 1.51 (1.49-1.53) | 1.42 (1.40-1.44) | 1.40 (1.39-1.42) |
| 5 least deprived | 142,315 (83,545) | 68.6 | 1.65 (1.63-1.67) | 1.54 (1.53-1.56) | 1.52 (1.50-1.54) |
| **†Setting** | | | | | |
| Urban city or town (ref) | 362,390 (186,715) | 61.2 | 1.00 | 1.00 | 1.00 |
| Urban conurbation | 138,875 (64,560) | 55.9 | 0.96 (0.95-0.97) | 1.02 (1.01-1.03) | 1.01 (1.00-1.03) |
| Rural town or village | 178,145 (98,170) | 64.7 | 1.08 (1.07-1.09) | 1.00 (0.99-1.01) | 1.00 (0.99-1.01) |
| **Kidney disease subgroup** | | | | | |
| CKD3a (ref) | 418,095 (228,710) | 62.5 | 1.00 | 1.00 | 1.00 |
| CKD3b | 193,475 (92,595) | 59.3 | 0.91 (0.90-0.91) | 0.92 (0.92-0.93) | 0.92 (0.91-0.93) |
| CKD4–5 | 50,465 (19,400) | 55.2 | 0.83 (0.82-0.84) | 0.86 (0.85-0.87) | 0.86 (0.85-0.87) |
| RRT (dialysis) | 4,595 (1,715) | 50.0 | 1.10 (1.05-1.15) | 1.19 (1.13-1.24) | 1.18 (1.12-1.23) |
| RRT (transplant) | 12,770 (7,025) | 58.9 | 2.71 (2.62-2.80) | 2.96 (2.86-3.06) | 3.08 (2.98-3.18) |
| **Primary care coding of kidney disease** | | | | | |
| CKD diagnostic code | 424,480 (217,290) | 61.2 | 1.00 (1.00-1.01) | 1.02 (1.01-1.02) | 1.04 (1.04-1.05) |
| **Risk group (occupation/access)** | | | | | |
| *†Care home resident | 43,385 (11,695) | 49.5 | 0.67 (0.65-0.68) | 0.73 (0.72-0.74) | 0.79 (0.77-0.80) |
| *†Health/social care worker | 1,095 (645) | 59.6 | 1.46 (1.35-1.58) | 1.50 (1.39-1.63) | 1.46 (1.35-1.58) |
| †Housebound | 49,420 (14,125) | 43.3 | 0.57 (0.56-0.58) | 0.60 (0.59-0.61) | 0.61 (0.60-0.62) |
| †End of life care | 35,765 (9,855) | 51.4 | 0.83 (0.82-0.85) | 0.88 (0.86-0.89) | 0.88 (0.86-0.90) |
| **Risk group (clinical)** | | | | | |
| †Prior SARS-CoV-2 | 19,170 (6,395) | 51.5 | 0.86 (0.84-0.89) | 0.92 (0.90-0.94) | 0.93 (0.90-0.95) |
| †Immunosuppression | 80,025 (38,925) | 56.7 | 1.32 (1.30-1.33) | 1.26 (1.24-1.28) | 1.21 (1.20-1.23) |
| Moderate/severe obesity | 55,040 (26,605) | 55.1 | 0.84 (0.83-0.85) | 0.89 (0.88-0.90) | 0.92 (0.91-0.93) |
| Diabetes | 190,140 (89,055) | 57.2 | 0.87 (0.87-0.88) | 0.92 (0.91-0.93) | 0.93 (0.92-0.94) |
| CRD (inc. asthma) | 151,400 (73,560) | 59.6 | 0.97 (0.96-0.98) | 1.00 (0.99-1.01) | 1.01 (1.00-1.02) |
| Chronic heart disease | 340,605 (167,470) | 60.9 | 0.97 (0.97-0.98) | 0.96 (0.95-0.97) | 0.97 (0.97-0.98) |
| Chronic liver disease | 18,715 (9,190) | 58.3 | 1.01 (0.99-1.03) | 1.03 (1.01-1.05) | 1.03 (1.01-1.05) |
| Asplenia | 5,930 (3,175) | 63.2 | 1.12 (1.08-1.16) | 1.09 (1.05-1.12) | 1.08 (1.05-1.12) |
| Cancer (non-haematologic) | 128,475 (66,290) | 64.3 | 1.10 (1.09-1.11) | 1.07 (1.06-1.08) | 1.07 (1.06-1.08) |
| †Haematologic cancer | 19,990 (10,400) | 64.4 | 1.47 (1.44-1.50) | 1.23 (1.20-1.26) | 1.33 (1.30-1.36) |
| Organ transplant (non-kidney) | 2,170 (1,210) | 62.3 | 1.81 (1.71-1.92) | 1.82 (1.72-1.93) | 2.41 (2.28-2.56) |
| CND (inc. learning disability) | 131,900 (55,625) | 56.2 | 0.86 (0.85-0.87) | 0.87 (0.86-0.88) | 0.88 (0.87-0.89) |
| Severe mental illness | 9,190 (3,195) | 44.0 | 0.67 (0.65-0.69) | 0.72 (0.69-0.74) | 0.74 (0.71-0.76) |
| CEV (other) | 3,990 (2,045) | 61.2 | 1.04 (1.00-1.09) | 1.10 (1.05-1.15) | 1.05 (1.01-1.10) |

Cumulative coverage was determined based on Kaplan-Meier estimates, censoring on death, deregistration, or 11$^{th}$ May 2022. * variable included in all minimally adjusted models; † variable included in all partially adjusted models. CI, confidence interval; CKD, chronic kidney disease; HR, hazard ratio; IMD, index of multiple deprivation; OR, odds ratio; RRT, renal replacement therapy.

**Supplementary Table 6. Cox proportional hazards models for factors associated with completion of a 3-dose vaccination series in kidney disease subgroups.**

| | CKD3a | | | CKD3b | | | CKD4–5 | | | RRT (dialysis) | | | RRT (transplant) | | |
|---|---|---|---|---|---|---|---|---|---|---|---|---|---|---|---|
| **Variable** | **N (n events)** | **Cov. (%)** | **HR (95% CI)** | **N (n events)** | **Cov. (%)** | **HR (95% CI)** | **N (n events)** | **Cov. (%)** | **HR (95% CI)** | **N (n events)** | **Cov. (%)** | **HR (95% CI)** | **N (n events)** | **Cov. (%)** | **HR (95% CI)** |
| **Age** | | | | | | | | | | | | | | | |
| 16–64 (ref) | 6,775 (2,000) | 31.0 | 1.00 | 1,845 (585) | 34.4 | 1.00 | 800 (255) | 35.4 | 1.00 | 1,300 (440) | 37.3 | 1.00 | 9,295 (4,730) | 53.3 | 1.00 |
| 65–69 | 4,705 (1,630) | 37.2 | 1.21 (1.13–1.29) | 1,395 (515) | 41.3 | 1.21 (1.08–1.37) | 435 (165) | 43.7 | 1.36 (1.11–1.65) | 245 (85) | 40.8 | 1.22 (0.97–1.54) | 1,425 (945) | 72.1 | 1.60 (1.49–1.72) |
| 70–74 | 8,730 (3,790) | 47.9 | 1.61 (1.52–1.70) | 2,815 (1,105) | 46.0 | 1.27 (1.15–1.41) | 825 (305) | 48.1 | 1.37 (1.16–1.62) | 245 (90) | 48.1 | 1.35 (1.07–1.70) | 1,180 (800) | 75.9 | 1.67 (1.55–1.81) |
| 75–79 | 124,030 (74,145) | 63.9 | 2.58 (2.46–2.71) | 38,245 (20,495) | 59.9 | 1.97 (1.81–2.15) | 8,410 (3,765) | 56.3 | 1.76 (1.54–2.02) | 1,220 (495) | 55.6 | 1.45 (1.19–1.75) | 620 (410) | 76.2 | 1.67 (1.51–1.85) |
| 80+ | 273,855 (147,145) | 63.7 | 2.79 (2.66–2.92) | 149,170 (69,900) | 59.9 | 2.12 (1.95–2.30) | 40,000 (14,910) | 55.7 | 1.82 (1.59–2.08) | 1,590 (605) | 58.1 | 1.62 (1.33–1.96) | 250 (140) | 73.5 | 1.68 (1.42–1.99) |
| **Sex** | | | | | | | | | | | | | | | |
| Female (ref) | 241,015 (127,790) | 60.2 | 1.00 | 112,810 (51,900) | 56.4 | 1.00 | 27,035 (9,830) | 51.4 | 1.00 | 1,710 (585) | 44.9 | 1.00 | 4,925 (2,825) | 60.8 | 1.00 |
| Male | 177,080 (100,920) | 65.8 | 1.15 (1.14–1.16) | 80,665 (40,695) | 63.3 | 1.19 (1.18–1.21) | 23,430 (9,565) | 59.8 | 1.24 (1.20–1.27) | 2,885 (1,130) | 52.8 | 1.19 (1.07–1.31) | 7,845 (4,200) | 57.7 | 0.92 (0.87–0.96) |
| **Ethnicity** | | | | | | | | | | | | | | | |
| White (ref) | 399,635 (223,240) | 63.9 | 1.00 | 184,715 (90,355) | 60.6 | 1.00 | 47,335 (18,735) | 57.0 | 1.00 | 3,755 (1,480) | 54.0 | 1.00 | 10,390 (6,240) | 64.3 | 1.00 |
| Mixed | 1,620 (550) | 38.1 | 0.54 (0.50–0.59) | 710 (210) | 35.5 | 0.51 (0.45–0.58) | 215 (55) | 33.3 | 0.59 (0.45–0.77) | 60 (15) | 20.0 | 0.59 (0.35–0.97) | 140 (45) | 30.8 | 0.49 (0.36–0.65) |
| Asian or Asian British | 9,955 (2,965) | 33.8 | 0.46 (0.45–0.48) | 4,920 (1,220) | 30.5 | 0.45 (0.43–0.48) | 1,820 (360) | 28.1 | 0.46 (0.41–0.51) | 490 (140) | 35.6 | 0.59 (0.49–0.71) | 1,510 (515) | 36.1 | 0.51 (0.47–0.56) |
| Black or Black British | 4,630 (1,020) | 24.4 | 0.34 (0.32–0.36) | 2,155 (455) | 24.9 | 0.37 (0.33–0.40) | 780 (150) | 23.1 | 0.39 (0.33–0.46) | 235 (60) | 32.6 | 0.53 (0.40–0.69) | 470 (120) | 27.3 | 0.38 (0.32–0.46) |
| Other ethnic groups | 2,255 (935) | 47.4 | 0.70 (0.66–0.75) | 980 (355) | 44.2 | 0.70 (0.63–0.78) | 315 (100) | 42.8 | 0.75 (0.61–0.91) | 55 (15) | 20.0 | 0.59 (0.35–0.98) | 260 (105) | 41.7 | 0.63 (0.51–0.76) |
| **IMD** | | | | | | | | | | | | | | | |
| 1 most deprived (ref) | 60,420 (26,045) | 50.4 | 1.00 | 30,950 (11,780) | 48.2 | 1.00 | 8,705 (2,590) | 43.7 | 1.00 | 1,015 (285) | 36.5 | 1.00 | 2,600 (985) | 40.3 | 1.00 |
| 2 | 74,870 (37,585) | 58.0 | 1.19 (1.17–1.21) | 36,495 (16,045) | 55.0 | 1.17 (1.14–1.20) | 9,935 (3,525) | 51.5 | 1.19 (1.13–1.25) | 1,005 (355) | 47.4 | 1.40 (1.19–1.64) | 2,665 (1,290) | 51.5 | 1.37 (1.26–1.49) |
| 3 | 95,730 (52,840) | 63.0 | 1.31 (1.29–1.33) | 44,525 (21,715) | 60.2 | 1.30 (1.27–1.33) | 11,530 (4,530) | 56.3 | 1.30 (1.23–1.37) | 980 (395) | 53.1 | 1.54 (1.31–1.81) | 2,760 (1,625) | 63.0 | 1.69 (1.55–1.83) |
| 4 | 95,975 (55,950) | 66.1 | 1.41 (1.39–1.43) | 42,745 (21,840) | 62.8 | 1.39 (1.36–1.42) | 10,810 (4,560) | 59.3 | 1.40 (1.33–1.47) | 845 (355) | 55.4 | 1.61 (1.37–1.91) | 2,520 (1,605) | 67.9 | 1.86 (1.71–2.02) |
| 5 least deprived | 91,100 (56,295) | 69.7 | 1.53 (1.51–1.55) | 38,755 (21,215) | 66.6 | 1.51 (1.48–1.55) | 9,485 (4,195) | 62.8 | 1.49 (1.42–1.57) | 745 (320) | 57.7 | 1.73 (1.46–2.05) | 2,225 (1,525) | 72.7 | 2.05 (1.88–2.23) |
| **Setting** | | | | | | | | | | | | | | | |
| Urban city or town (ref) | 222,365 (121,865) | 62.7 | 1.00 | 103,970 (50,030) | 59.6 | 1.00 | 27,115 (10,385) | 55.1 | 1.00 | 2,320 (805) | 46.6 | 1.00 | 6,615 (3,630) | 58.8 | 1.00 |
| Urban conurbation | 83,815 (41,785) | 57.7 | 1.02 (1.01–1.04) | 39,355 (16,900) | 53.9 | 1.01 (0.99–1.03) | 10,895 (3,770) | 50.1 | 1.04 (0.99–1.09) | 1,320 (480) | 47.4 | 1.08 (0.92–1.26) | 3,495 (1,625) | 49.5 | 1.05 (0.97–1.14) |
| Rural town or village | 111,915 (65,065) | 65.8 | 1.00 (0.99–1.01) | 50,150 (25,665) | 62.7 | 0.99 (0.98–1.01) | 12,460 (5,245) | 60.0 | 1.03 (0.99–1.06) | 960 (430) | 59.9 | 1.19 (1.06–1.35) | 2,660 (1,770) | 71.0 | 1.17 (1.10–1.23) |
| **Primary care coding of kidney disease** | | | | | | | | | | | | | | | |
| CKD diagnostic code | 214,160 (118,570) | 63.2 | 1.03 (1.02–1.03) | 151,790 (74,260) | 59.9 | 1.07 (1.05–1.08) | 44,890 (17,535) | 55.6 | 1.06 (1.01–1.12) | 3,985 (1,485) | 50.7 | 0.98 (0.85–1.14) | 9,655 (5,435) | 60.6 | 1.13 (1.07–1.19) |
| **Risk group (occupation/access)** | | | | | | | | | | | | | | | |
| Care home resident | 23,700 (6,855) | 49.5 | 0.72 (0.70–0.74) | 14,845 (3,800) | 49.4 | 0.76 (0.74–0.79) | 4,605 (995) | 49.9 | 0.86 (0.80–0.92) | 165 (30) | 22.2 | 0.64 (0.44–0.94) | 65 (20) | 33.3 | 0.46 (0.30–0.72) |
| Health/social care worker | 565 (325) | 58.1 | 1.53 (1.37–1.70) | 120 (70) | 50.0 | 1.36 (1.08–1.73) | [R] | [R] | [R] | [R] | [R] | [R] | 355 (225) | 61.1 | 1.31 (1.15–1.50) |
| Housebound | 24,345 (7,520) | 44.2 | 0.59 (0.58–0.60) | 17,740 (4,835) | 42.1 | 0.59 (0.58–0.61) | 6,345 (1,320) | 38.8 | 0.60 (0.57–0.64) | 375 (100) | 44.9 | 0.88 (0.72–1.08) | 615 (345) | 61.5 | 0.95 (0.85–1.06) |
| End of life care | 17,935 (5,395) | 52.4 | 0.87 (0.84–0.89) | 12,290 (3,240) | 50.5 | 0.89 (0.86–0.92) | 4,995 (1,035) | 48.5 | 0.92 (0.86–0.98) | 365 (90) | 43.5 | 0.77 (0.63–0.96) | 180 (90) | 68.3 | 1.26 (1.02–1.56) |
| **Risk group (clinical)** | | | | | | | | | | | | | | | |
| Prior SARS-CoV-2 | 10,080 (3,705) | 53.2 | 0.92 (0.89–0.95) | 6,040 (1,840) | 50.4 | 0.92 (0.87–0.96) | 2,065 (485) | 47.0 | 0.91 (0.83–1.00) | 475 (140) | 41.7 | 0.85 (0.71–1.02) | 515 (230) | 46.7 | 0.88 (0.77–1.00) |
| Immunosuppression | 43,575 (21,400) | 55.8 | 1.22 (1.19–1.24) | 18,245 (8,595) | 58.1 | 1.26 (1.22–1.29) | 5,525 (2,200) | 55.9 | 1.34 (1.26–1.42) | 2,065 (750) | 44.2 | 1.05 (0.88–1.25) | 10,615 (5,975) | 60.3 | 1.17 (1.10–1.26) |
| Moderate/severe obesity | 31,670 (16,170) | 56.1 | 0.90 (0.89–0.92) | 16,970 (7,815) | 54.1 | 0.91 (0.89–0.93) | 4,880 (1,865) | 50.4 | 0.92 (0.87–0.96) | 360 (130) | 45.9 | 0.99 (0.83–1.19) | 1,160 (630) | 56.7 | 1.03 (0.95–1.12) |
| Diabetes | 100,110 (50,045) | 58.2 | 0.91 (0.90–0.92) | 62,990 (28,520) | 56.6 | 0.94 (0.93–0.96) | 21,315 (7,790) | 53.1 | 0.95 (0.92–0.98) | 1,685 (555) | 48.2 | 0.96 (0.86–1.06) | 4,040 (2,145) | 58.4 | 1.03 (0.98–1.09) |
| CRD (inc. asthma) | 91,725 (47,415) | 60.9 | 1.00 (0.99–1.01) | 44,605 (20,325) | 58.2 | 1.01 (0.99–1.02) | 11,810 (4,285) | 54.3 | 1.01 (0.98–1.05) | 1,005 (355) | 50.1 | 1.08 (0.96–1.21) | 2,255 (1,175) | 56.1 | 0.93 (0.87–0.99) |
| Chronic heart disease | 195,385 (103,225) | 62.5 | 0.96 (0.95–0.97) | 107,965 (49,800) | 59.2 | 0.97 (0.96–0.99) | 30,860 (11,355) | 55.5 | 0.98 (0.95–1.01) | 2,535 (910) | 51.4 | 0.98 (0.89–1.08) | 3,860 (2,185) | 63.3 | 1.05 (0.99–1.10) |
| Chronic liver disease | 11,090 (5,770) | 59.4 | 1.05 (1.02–1.08) | 5,290 (2,410) | 56.3 | 1.02 (0.98–1.06) | 1,505 (585) | 55.0 | 1.06 (0.98–1.16) | 240 (85) | 44.4 | 1.18 (0.94–1.47) | 585 (340) | 60.9 | 1.22 (1.09–1.36) |
| Asplenia | 3,650 (2,075) | 64.5 | 1.09 (1.04–1.14) | 1,575 (795) | 61.1 | 1.10 (1.02–1.18) | 455 (180) | 55.5 | 1.08 (0.93–1.25) | 80 (35) | 50.0 | 1.51 (1.07–2.14) | 170 (90) | 56.2 | 0.98 (0.80–1.20) |
| Cancer (non-haematologic) | 77,375 (42,645) | 65.7 | 1.07 (1.06–1.09) | 38,315 (18,430) | 62.4 | 1.07 (1.05–1.09) | 11,020 (4,295) | 59.0 | 1.08 (1.05–1.12) | 805 (320) | 54.7 | 1.05 (0.93–1.19) | 960 (600) | 70.7 | 1.16 (1.07–1.26) |
| Haematologic cancer | 11,885 (6,460) | 64.3 | 1.33 (1.29–1.36) | 5,705 (2,895) | 64.8 | 1.33 (1.27–1.39) | 1,845 (765) | 61.5 | 1.32 (1.20–1.44) | 310 (135) | 58.6 | 1.70 (1.40–2.06) | 240 (145) | 63.6 | 1.06 (0.90–1.25) |
| Organ transplant (non-kidney) | 1,195 (700) | 62.4 | 2.57 (2.38–2.77) | 685 (365) | 60.8 | 2.26 (2.03–2.52) | 215 (110) | 57.2 | 1.97 (1.62–2.40) | 75 (35) | 42.9 | 1.72 (1.23–2.41) | [N] | [N] | [N] |
| CND (inc. learning disability) | 77,535 (35,190) | 57.6 | 0.87 (0.86–0.88) | 41,125 (16,030) | 54.3 | 0.87 (0.85–0.88) | 11,295 (3,520) | 51.6 | 0.90 (0.87–0.94) | 765 (235) | 44.6 | 0.87 (0.76–1.01) | 1,185 (645) | 60.1 | 1.01 (0.93–1.09) |
| Severe mental illness | 5,405 (1,945) | 43.8 | 0.70 (0.67–0.74) | 2,700 (895) | 43.6 | 0.74 (0.70–0.79) | 835 (250) | 44.9 | 0.85 (0.75–0.96) | 80 (20) | 28.6 | 0.71 (0.46–1.09) | 170 (85) | 50.0 | 0.99 (0.79–1.22) |
| CEV (other) | 2,370 (1,280) | 61.8 | 1.09 (1.03–1.15) | 1,170 (575) | 60.5 | 1.15 (1.06–1.25) | 450 (190) | 55.3 | 1.10 (0.96–1.27) | [N] | [N] | [N] | [N] | [N] | [N] |

Individuals were included in the analysis if they were ≥75 years of age at baseline, care home residents, transplant recipients, or had a history of haematologic malignancy or immunosuppression. Counts are rounded to the nearest 5. Cumulative coverage was determined based on Kaplan-Meier estimates, censoring on death, deregistration, or 11th May 2022. Hazard ratios and 95% confidence intervals were derived from partially adjusted models that included age, care home residence, health and social care worker status, housebound status, receipt of end-of-life care, setting (urban/rural), sex, ethnicity, IMD quintile, prior SARS-CoV-2 infection, immunosuppression, and haematologic cancer. Data are redacted [R] for any rows in which there were >0 and ≤10 event or non-event counts after rounding to the nearest 5, with non-event counts calculated among uncensored individuals. CEV, clinically extremely vulnerable; CKD, chronic kidney disease; CND, chronic neurological disease; IMD, index of multiple deprivation; [N], covariate absent in all individuals by definition; [R] redacted; RRT, renal replacement therapy.
